# Comprehensive genome-wide association study of different forms of hernia identifies more than 80 associated loci

**DOI:** 10.1101/2021.04.27.21256188

**Authors:** João Fadista, Line Skotte, Juha Karjalainen, Erik Abner, Erik Sørensen, Henrik Ullum, Thomas Werge, iPSYCH-group, Tõnu Esko, Lili Milani, Aarno Palotie, Mark Daly, FinnGen, Mads Melbye, Bjarke Feenstra, Frank Geller

## Abstract

Hernias are characterized by protrusion of an organ or tissue through its surrounding cavity and often require surgical repair. We identified 65,492 cases for five hernia types in the UK Biobank and performed genome-wide association study scans for these five types and two combined groups. The results showed associated variants in all scans. Inguinal hernia had the most associations and we conducted a replication study with 23,803 additional cases from four study groups giving 84 independently associated variants. Identified variants from all scans were collapsed into 81 independent loci. Further testing showed that 26 loci were associated with more than one hernia type, suggesting substantial overlap between the underlying genetic mechanisms. Pathway analysis identified several genes with a strong link to collagen and/or elastin (*ADAMTS6, ADAMTS16, ADAMTSL3, LOX, ELN*) in the vicinity of associated loci for inguinal hernia, which substantiates an essential role of connective tissue morphology.

## Introduction

A hernia is an outpouching of tissue through a preformed or secondarily established fissure^1^. Most hernias occur in the abdominal wall, only diaphragmatic hernias present with a defect of the diaphragm resulting in abdominal tissue entering the chest cavity. A Swedish study on the familial risk of different hernias, including inguinal, umbilical, femoral and incisional hernias, identified increased sibling risks not only for pairs with the same form of hernia, also for several combinations of discordant hernias^2^. They concluded that genetic factors play a role in the etiology of hernias. The fact that increased risks were seen across different forms of hernia supports the hypothesis that there might be genetic risk factors contributing to multiple forms of hernia and combined analyses have potential to identify additional associated loci. Higher BMI is associated with increased and decreased risk of diaphragmatic hernia^3^ and inguinal hernia^4^, respectively. On the other hand, inguinal hernia and gastroesophagal reflux (a frequent consequence of diaphragmatic hernia) have been associated with psychiatric disorders later in life, including depression and schizophrenia^5,6^. Comprehensive GWAS data on hernias are needed to investigate potential shared genetic susceptibility between hernias and these and other traits. So far, the only genome-wide association study on hernias was on inguinal hernias and identified four risk loci^7^.

Starting with data from the UK Biobank^8^, we investigate five different forms of hernia classified according to ICD 10 codes by their location as inguinal (K40), femoral (K41), umbilical (K42), and diaphragmatic (K44), or by their origin in previous injury of the abdominal wall (ventral hernia, K43). To maximize power, we combine information about hernia from hospital records (surgery codes, ICD9- and ICD10-codes) and interview data (surgery and non-cancer illness). We also study two combined groups: any hernia and any hernia excluding individuals who only had diaphragmatic hernia, as diaphragmatic hernia is special by its location and the substantially lower rate of surgery-confirmed cases. Variants associated with inguinal hernia are put forward to *in silico* replication in four additional study groups.

## Results

### Genome-wide association scans in the UK Biobank

**Figure 1** presents the design of the study, including sample sizes for the different forms of hernia in the UK Biobank and the set-up of the inguinal hernia replication study. We performed genome-wide association studies (GWAS) scans of all individuals with British ancestry confirmed by genetic principal components analysis for the 7 hernia groups with SAIGE^9^, a method that adjusts for case-control imbalance and relatedness amongst individuals. Some individuals had more than one form of hernia and the correlation between the different hernia groups (**Supplementary Table 1**) ranged from *r* = 0.006 (95%-CI: 0.003 – 0.009) between diaphragmatic and femoral to *r* = 0.133 (95%-CI: 0.130 – 0.136) between umbilical and ventral. The combined groups were strongly correlated (all *r* > 0.6) with the large hernia groups they included (**Supplementary Table 1**). We performed the analyses for the full group of cases and controls (*all* analysis), as well as for the subgroups of only male or female cases and controls (male / female analysis) For all GWAS scans we used individuals without any form of hernia as controls, *P* values were adjusted by genomic control, ranging from 1.00 (femoral) to 1.18 (any hernia, **Supplementary Table 1**). All 7 scans for the different hernia groups identified genetic variants at genome-wide significance. **Figure 2** shows Manhattan plots for the 7 analyzed phenotypes). We did a conditional and joined analysis of the initial SAIGE results with GCTA-COJO^10^ to also investigate multiple hits per locus, and we observed between two and 55 independently associated variants per phenotype at *P* < 5 × 10^−8^. (**Supplementary Table 2**). There was overlap between the identified loci per hernia type and the combined groups identified additional loci. A genetic link between the different forms of hernia was further supported by genetic correlation analysis of the five scans for specific forms of hernia, where we observed significantly increased *rg*’s for all combinations ranging from 0.24 (95%-CI: 0.17-0.31) between inguinal and diaphragmatic to 0.76 (95%-CI: 0.56-0.95) between femoral and ventral (**Supplementary Table 1**).

**Figure 1:**
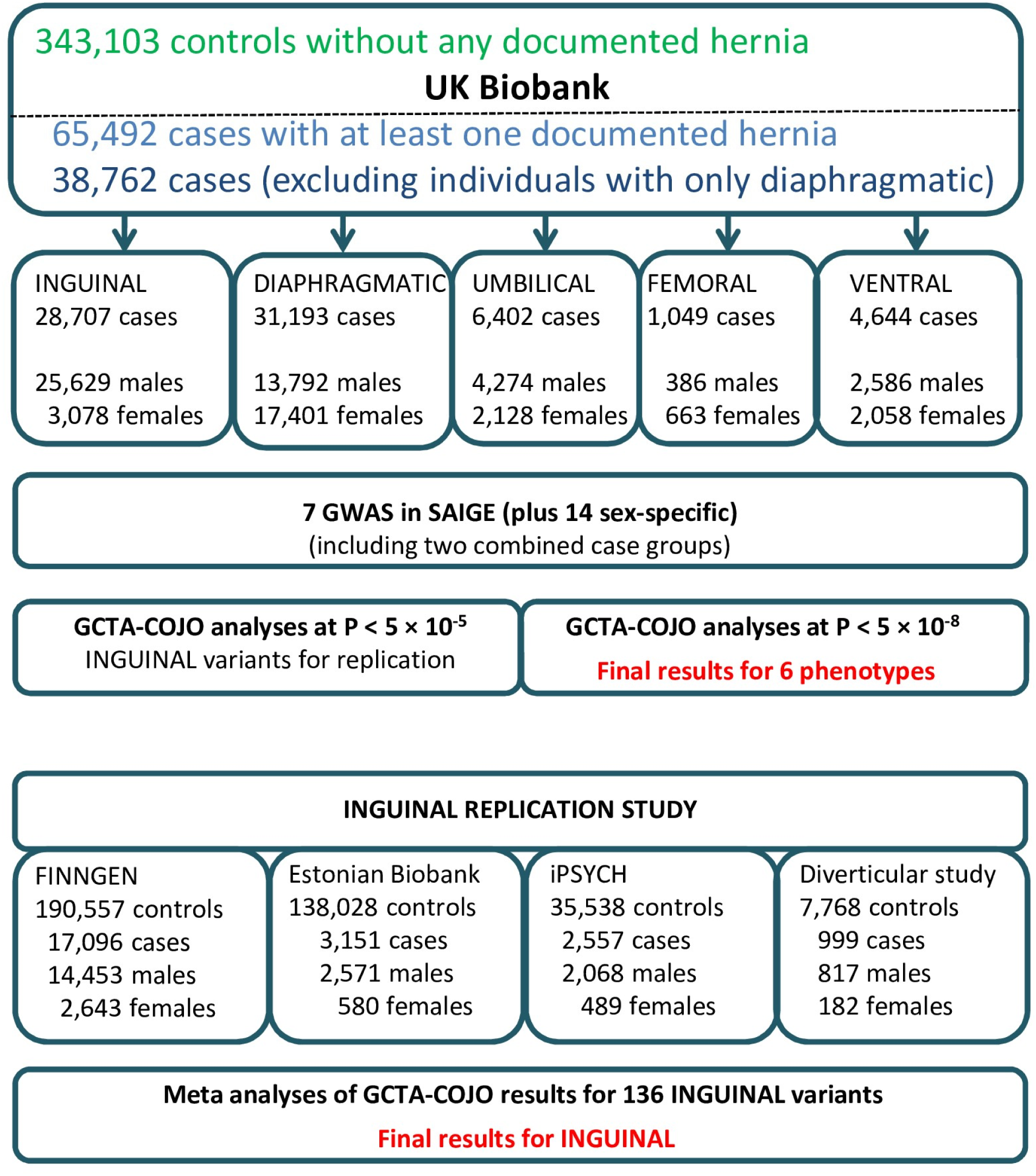
Study design and sample sizes for the study groups.

**Figure 2:**
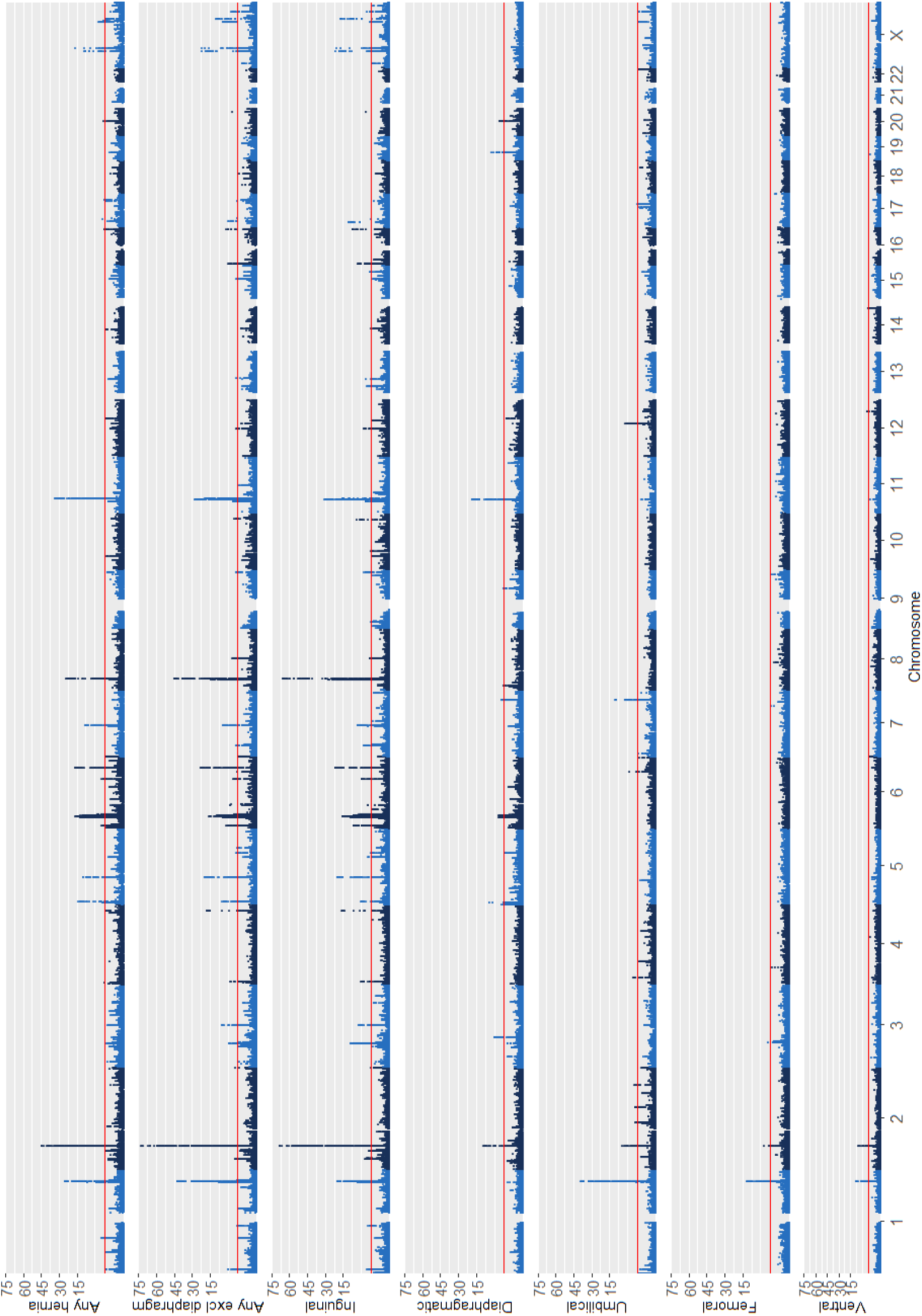
Manhattan plots for the 7 analyzed hernia groups, displaying –log10 (*P* values) from the SAIGE analyses over the chromosomes. The scale for the y-axis is shrinked by a factor of 5 for values above 15. The red line indicates the *P* threshold of 5 × 10^−8^.

The sex-specific analyses mainly identified associated variants at *P* < 5 × 10^−8^ in the male analyses and most overlapped with loci identified in the *all* analyses for these phenotypes (**Supplementary Table 2**). Furthermore, the analyses showed that the most significantly associated loci can differ between males and females (**Supplementary Note)**.

### Replication of variants associated with inguinal hernia

For inguinal hernia, we were able to perform *in silico* replication in the FINNGEN project, the Estonian Biobank and two Danish cohorts, providing replication findings for almost 24,000 cases and more than 370,000 controls (**Figure 1**).The GCTA-COJO analyses identified several regions with multiple associated variants. Therefore, we used the results from GCTA-COJO analyses of inguinal hernia at *P* < 1 × 10^−5^ as starting point and identified these variants or surrogate SNPs with an *r*^*2*^ > 0.8 in the four replication groups. Of the 136 investigated variants, only three had no information in the replication groups, while two more were missing in both FINNGEN and Estonian Biobank. After performing GCTA-COJO analyses in the replication groups, the results were meta-analyzed with METAL^11^. The number of variants reaching genome-wide significance increased to 84 (from 53 in UK Biobank alone) in all cases, to 70 (48) in males and to 7 (4) in females (**Supplementary Table 3**). As expected, there was a large overlap between the *all* and the sex-specific findings, and some of the hits were already identified in the UK Biobank larger groups combining any hernia or any hernia excluding diaphragmatic.

### Summary of all identified loci

The loci identified for the different hernia groups partially overlapped, so we decided to collapse loci by binning variants with an *r*^2^ > 0.6 under a key SNP with the lowest *P* value in any of the UK Biobank GCTA-COJO analyses. For loci we chose a broad definition of 500 kb to the next associated key SNP and for key SNPs with *P* < 1 × 10^−30^ the distance was increased to 1 Mb as changes in the conditional *P* values were observed even over these distances; the HLA region on chromosome 6 was considered one locus. We added new variants identified in the inguinal meta-analyses after the initial binning.

In total, the analyses of the *all* groups resulted in 81 collapsed loci with 114 distinct key SNPs, the sex-specific analyses added another 6 loci (5 in males) and 21 key SNPs (**Supplementary Table 3**). Again, the findings were concentrated in the groups including inguinal hernia and for 69 of the 81 loci at least one key SNP showed the lowest *P* value for inguinal hernia or one of the combined groups including inguinal hernia, the inguinal hernia replication study alone accounted for 22 of these loci. Up to 6 distinct key SNPs were identified per locus; sometimes multiple key SNPs for the same phenotype, sometimes there was variation regarding the associated phenotypes or sexes. The larger number of sex-specific findings in males was at least partly driven by the higher statistical power due to larger numbers of male cases, especially for inguinal hernia. Five key SNPs were identified in the HLA region and subsequent comparison with HLA-alleles showed that none of these SNPs was a best match for a known HLA-allele, only one HLA allele showed an *r*^*2*^ > 0.5 for a key SNP (HLA-B*8:01 with rs9281226, *r*^*2*^ = 0.67).

We investigated heterogeneity between the sexes for the associated variants identified at genome-wide significance by meta-analyzing the results for males and females with METAL^11^. The analysis was restricted to variants genome-wide significant for specific forms of hernias as the composition of the combined groups largely differed between the sexes (**Supplementary Table 1**). Among 110 variants tested, 50 variants showed no indication for heterogeneity (*I*^*2*^ = 0) so that analyzing both sexes together had increased statistical power substantially (**Supplementary Table 3**). For 22 (21 inguinal) variants a high degree of heterogeneity (*I*^*2*^ > 75) was observed. However, this did not necessarily mean that the association was confined to one sex: 10 variants showed no indication of association with inguinal hernia in females (effect in other direction or *P* > 0.2), but in the other 12 instances the analysis for the other sex reached at least nominal significance (range of higher sex-specific *P*: 1.6 × 10^−3^ to 1.9 × 10^−14^, for 10 of these variants the stronger effect estimates were observed in females).

### MultiPhen and variants with effects in opposite directions in different types of hernia

As some variants were associated with more than one hernia group and the two combined hernia groups only cover a small fraction of all combinations of hernia types, we decided to pinpoint the associated hernia types by running MultiPhen^12^ on the 114 associated key SNPs (**Supplementary Table 3**). For a specific variant, a hernia type was considered associated if it was associated at *P* < 0.01 and the results were summarized by locus, i.e. a hernia group was considered associated if any of the variants was associated. The summary of the associated hernia combinations for the 81 loci can be seen in **Figure 3**. A locus on chromosome 7q33 (key SNP rs12707188) displayed risk for umbilical and diaphragmatic hernia at genome-wide significance for two variants in LD (*r*^*2*^ = 0.76). Checking the effect in the other phenotype showed that the risk allele for umbilical hernia was protective for diaphragmatic and vice versa(**Supplementary Table 3**). The variants are located in *CALD1*, (calmodulin- and actin-binding protein), a gene important for smooth muscle and nonmuscle contraction. Studies on the molecular level are needed to investigate whether functional changes in this gene indeed mean opposite risk outcomes for the two forms of hernia.

**Figure 3:**
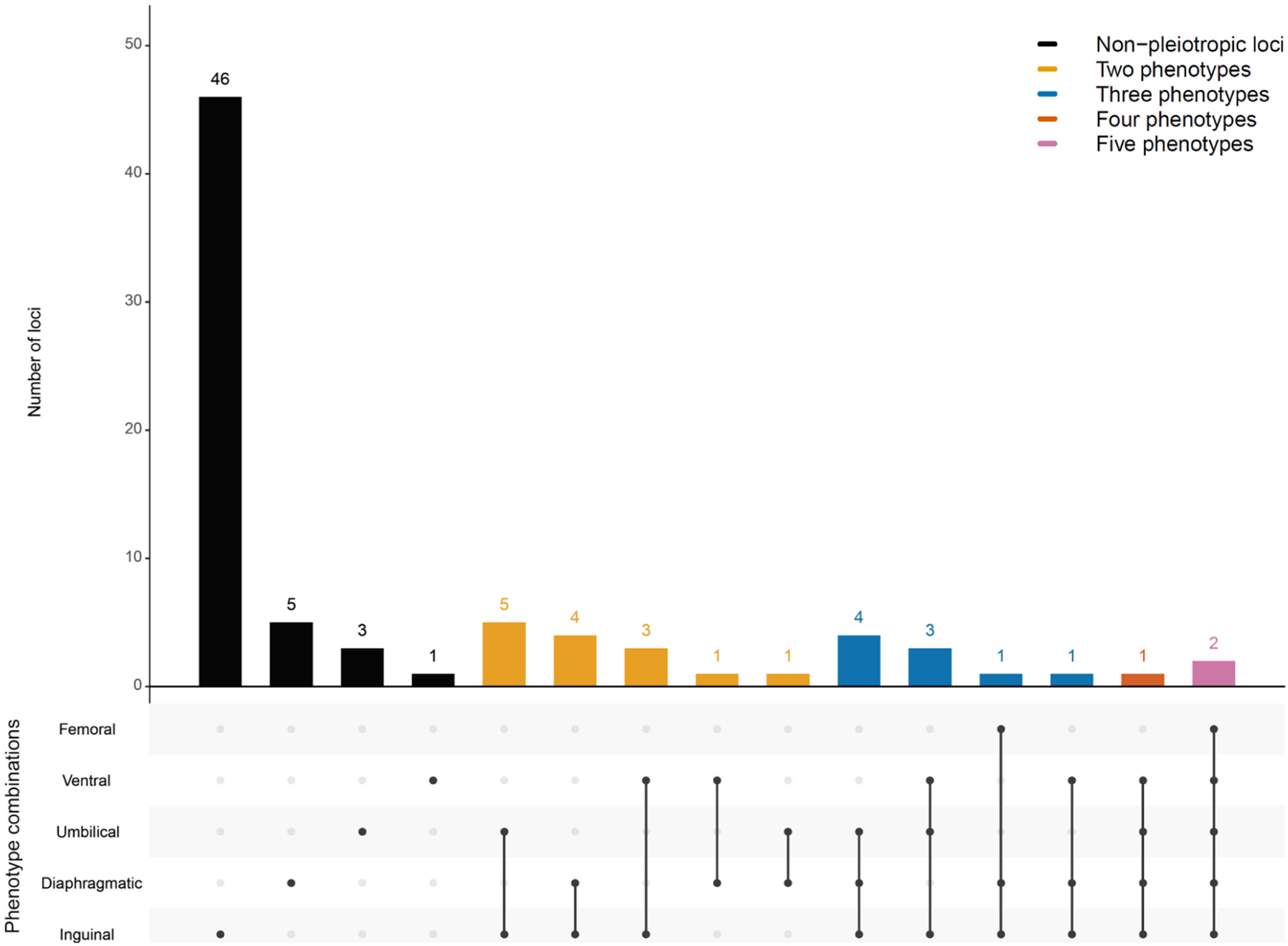
Distribution of the associated hernia traits (*P* < 0.01 in the MultiPhen analysis) for the 81 associated loci.

This finding lead us to investigate the potential of opposite effects on a broader scale by analyzing all variants with *P* < 0.001 for at least one of the 7 phenotypes under study with MultiPhen. We only considered variants outside the previously investigated collapsed loci and confirmed the initial results by linear regression in R^13^ on a subset of 335,258 unrelated individuals. Here we identified five new suggestive loci, four of which displayed effects going opposite directions, three times the hernia combination was inguinal and diaphragmatic and one time inguinal, umbilical and ventral (going the same direction as inguinal); the remaining locus had a main effect for umbilical seen at a lower level for diaphragmatic hernia. The five suggestive loci are displayed in **SupplementaryTable 3**.

To pinpoint genes and pathways responsible for the observed association findings and to investigate functional implications on a broader scale, we ran several pathway analysis tools on our results.

### MAGMA / FUMA

We applied gene prioritization by MAGMA^14^ as implemented in FUMA^15^; genes were considered associated if *P* < 2.55 × 10^−6^, the Bonferroni correction considering 19,622 genes analyzed. Genes were prioritized for all 7 analyzed phenotypes, with the largest numbers observed for inguinal hernia and the two combined groups (**Supplementary Table 4**).

FUMA expression results for 54 GTEx tissues were derived from MAGMA analysis. For inguinal hernia 17 tissues reached the group-wise significance (*P* < 9.26 × 10^−4^) including both adipose tissues, cultivated fibroblast cells and the esophagus muscle tissue, the findings in the two combined groups were very similar (**Supplementary Table 4**). Additional findings were seen for diaphragmatic hernia (artery aorta) und umbilical hernia (small intestine /terminal ileum and transverse colon).

### DEPICT

We also performed DEPICT^16^ analyses for each of the 7 analyzed phenotypes based on the associated variants with *P* < 1 × 10^−5^ as input. Thus, the gene prioritization is targeted on genes in the vicinity of these variants. Genes with a false discovery rate (FDR) below 5% were only found for inguinal hernia and the two combined groups (**Supplementary Table 5**). For inguinal hernia 64 of the 196 investigated genes had an FDR < 5% and we will take a closer look at these genes in the Discussion.

Similarly, in the DEPICT analyses of 10,968 genesets, we detected 451 sets with an FDR < 5% for inguinal hernia, and somewhat lower numbers for the two combined groups, whereas the other four hernia types had no findings (**Supplementary Table 5**). The geneset descriptions had many keywords like abnormal, morphology, morphogenesis, development, muscle, embryo, tissue and renal/kidney.

Enrichment of expression in particular tissues and cell types was studied by testing whether genes in associated regions were highly expressed in any of 209 Medical Subject Heading (MeSH) annotations in data from 37,427 microarrays(**Supplementary Table 5**). In total, 36 significant tissue or cell type annotations among the 209 analyzed categories were identified for any hernia (34), umbilical and any hernia excluding diaphragmatic (one each). The findings for any hernia substantiate trends observed in the analysis of inguinal hernia, the hits were mainly in the musculoskeletal, urogenital and cardiovascular systems, connective and muscle tissue cells and connective and membrane tissues(**Figure 4**).

**Figure 4:**
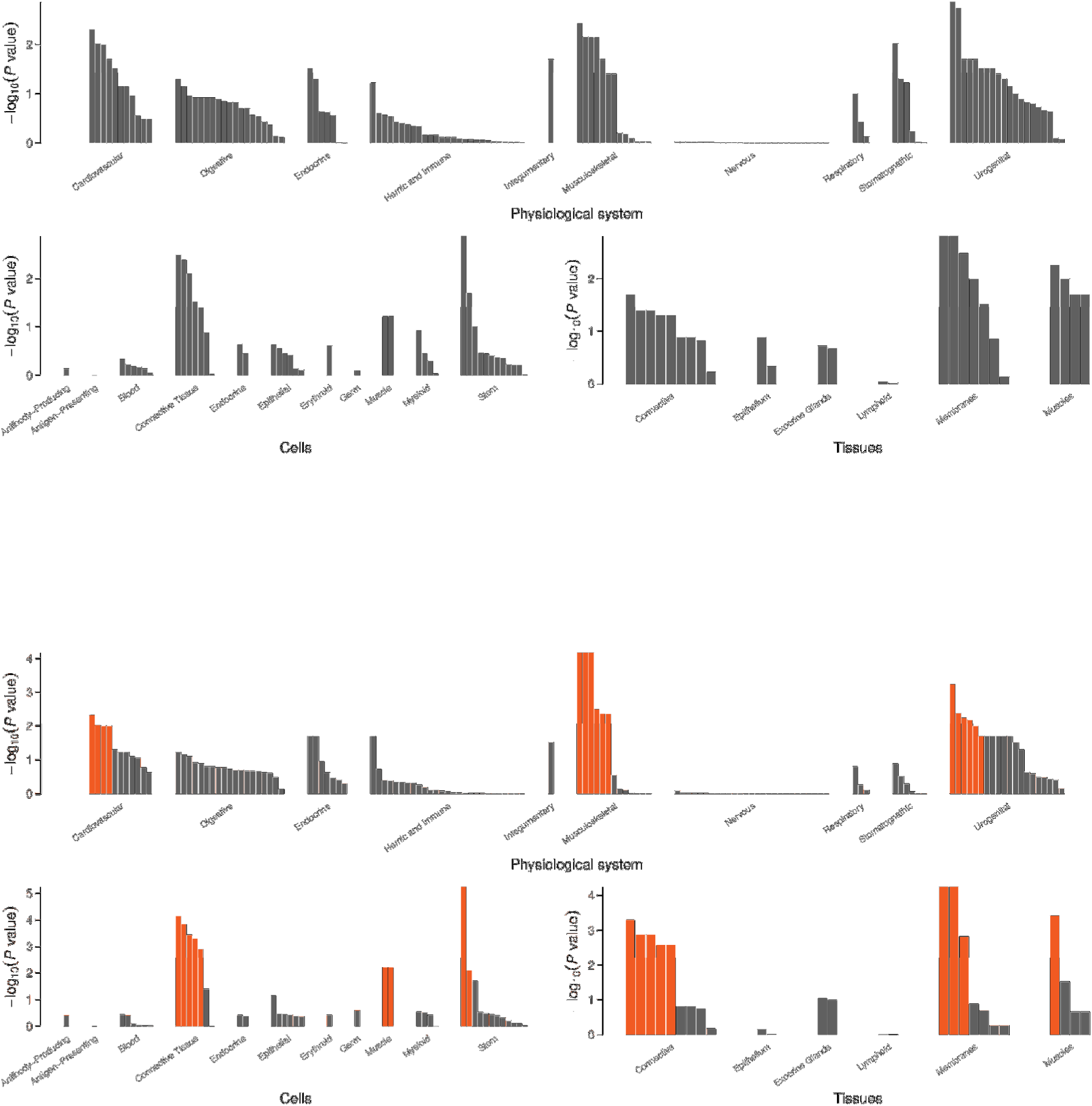
Plots showing the enrichment of loci associated with inguinal hernia (top) and any hernia (bottom) in physiological systems, cell types and tissues, colored annotations have FDR < 5%.

### GeneNetwork

GeneNetwork prioritizes genes based on expression data analysis specific for diseases based on human phenotype ontology (HPO) terms^17^. We compared results for the term “inguinal hernia” (HP: 0000023) in the 192 genes from the DEPICT gene prioritization for inguinal hernia with the referring DEPICT results(**Supplementary Table 5**). Nine of the genes showed significant results when adjusting for the 192 tests, 8 of which also had FDR < 5% in the DEPICT gene prioritization and 7 of the genes were in collapsed loci reaching genome-wide significance in our study.

### GARFIELD

We applied GARFIELD^18^ to assess the relative enrichment of phenotype–genotype associations to a generic regulatory annotation denoting open chromatin in 424 cell lines and primary cell types from ENCODE and Roadmap Epigenomics. The hotspots analyses of the DNaseI hypersensitive sites identified by far the most associations (for complete results for the 7 analyzed phenotypes see **Supplementary Table 6**). **Figure 5** illustrates the ubiquitous enrichment seen for inguinal hernia in an enrichment wheel plot. Many of the analyzed tissues were of fetal origin, and for several of them the enrichment was striking: at the level of *P* < 1 × 10^−5^, all 48 fetal muscle and all 11 fetal heart tissues had an empirical *P* < 1.18 × 10^−4^ (threshold adjusting for the 424 investigated cell types), another group with many enriched tissue probes was fibroblast, with all 19 tissues below the threshold.

**Figure 5:**
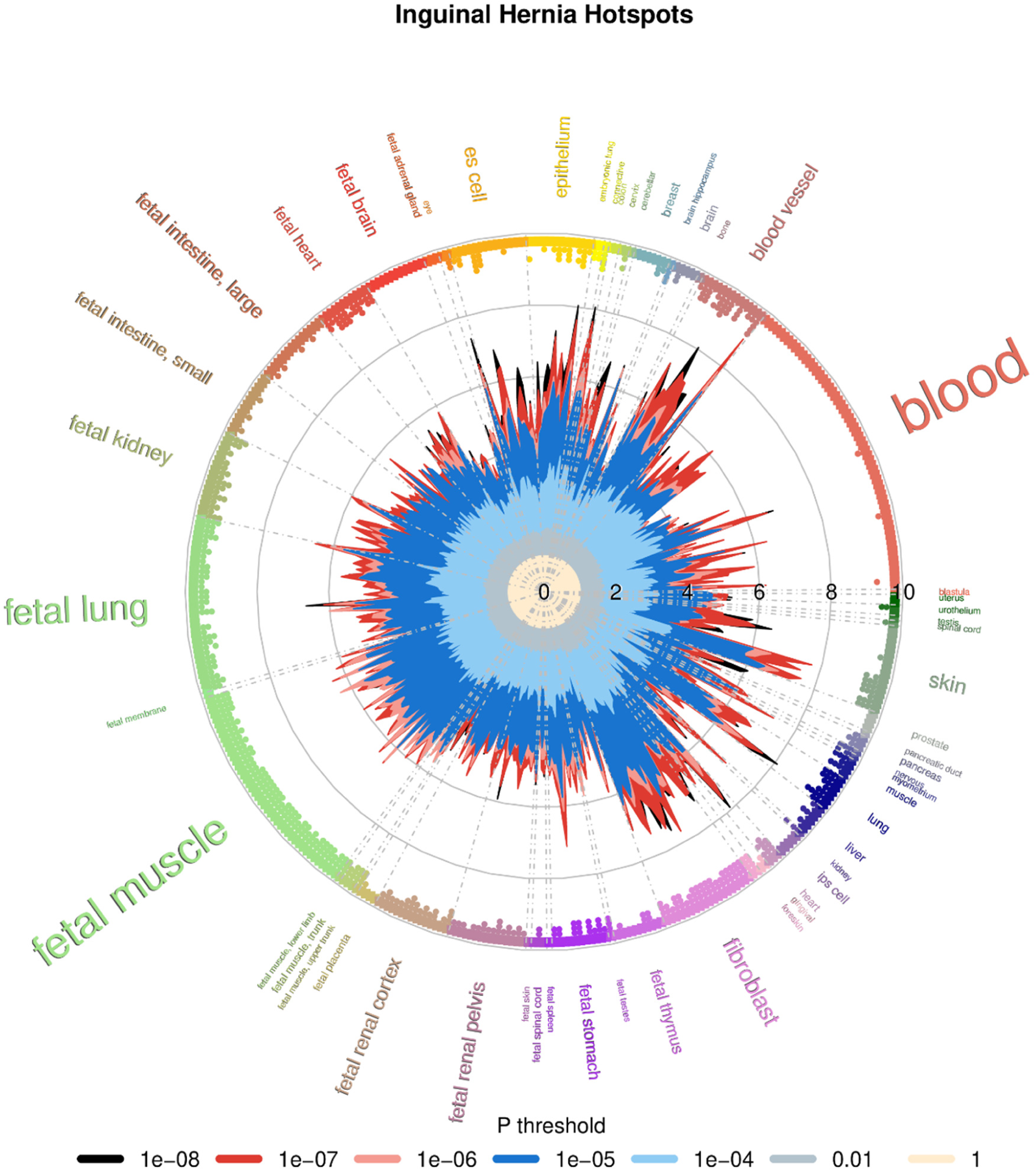
Enrichment wheel plot from the GARFIELD hotspots analysis for inguinal hernia. The radial spikes display the odds ratios for the tissues noted on the outside at 7 *P* value thresholds (color codes below the plot) for all ENCODE and Roadmap Epigenomics DHS cell lines. The dots just inside the outer circle denote significant (correcting for the number of effective annotations) GARFIELD enrichment separately for each of the five thresholds from 1 × 10^−4^ to 1 × 10^−8^.

### Gene expression in mice

We checked genes identified for hernia phenotypes in mice (Mammalian Phenotype Ontology Annotations: http://www.informatics.jax.org/vocab/mp_ontology) for overlap with genes annotated in the DEPICT analyses. In one mouse strain (C57BL/6), Efemp1(-/-) mice developed multiple large hernias including inguinal hernias, pelvic prolapse and protrusions of the xiphoid process, additional histological analysis revealed a marked reduction of elastic fibers in fascia^19^. In our analyses, the locus containing *EFEMP1* showed association with all five forms of hernia. However, the function of the encoded protein in humans is unknown.

### LD score regression / heritability

**Figure 6** shows the significant findings from bivariate LD score regression between the different forms of hernia and 245 traits(**Supplementary Table 7**), with the majority of findings for diaphragmatic hernia. In connection with the iPSYCH project^20^, we took a closer look at six psychiatric disorders and the finding for depressive symptoms and diaphragmatic hernia was also seen for major depressive disorder, both with diaphragmatic (*rg* = 0.23, 95%-CI: 0.10-0.35) and ventral hernia (*rg* = 0.45, 95%-CI: 0.19-0.71), all other correlations were not significant considering the 30 tests involved(**Supplementary Table 7**).

**Figure 6:**
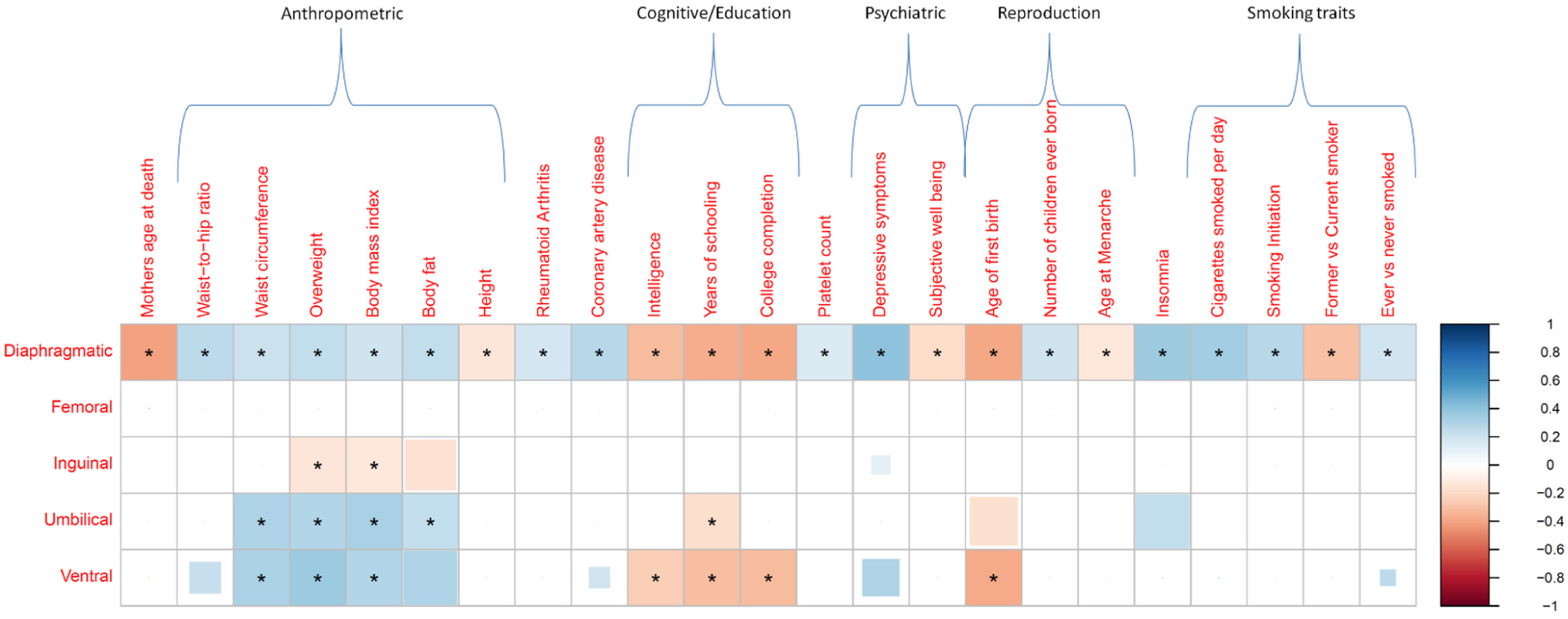
Pairwise genetic correlations estimated via bivariate LD score regression for 245 traits. Only correlations that were significant after correction for multiple testing in at least one form of hernia are shown (*P* < 2.04 × 10^−4^, indicated by an asterisk). Positive genetic correlations are shown in blue, negative ones in red for all combinations where *P* < 0.05, with the size of the colored squares indicating the significance-level (truncated at *P* < 2.04 × 10^−4^)

We estimated heritability for the five forms of hernia on the liability scale, for inguinal hernia it was 12.3% (95% CI: 9.8% - 14.7%), the other results were similar ranging from 9.0% (95% CI: 5.8% - 12.3%) for ventral hernia to 12.8% (95% CI: 9.5% - 16.1%) for umbilical hernia(**Supplementary Table 7**).

## Discussion

We performed GWAS scans on five different forms of hernia and two combined groups of any hernia and any hernia excluding diaphragmatic. Each individual scan resulted in at least two associated loci, but most associated variants were observed for inguinal hernia and the two combined groups, both including inguinal hernia. We were able to perform a replication study for inguinal hernia with almost 24,000 cases from four study groups. In the resulting meta-analysis, 84 variants reached genome-wide significance, confirming all 53 variants initially at that level in the UK Biobank plus an additional 31 which included 22 new loci. Summing up all variants reaching genome-wide significance in any of the analyses, we identified 114 independently associated variants in 81 loci. Sex-specific analyses identified an additional 21 associated variants (14 for males / 7 for females) which included 6 new loci. There are similarities between the different forms of hernia as all but diaphragmatic hernia are caused by weakness of the abdominal wall. Several variants were identified in more than one GWAS scan and there were additional variants identified through the two combined groups. In total, 39 of the 86 variants reaching genome-wide significance in the UK Biobank data alone displayed the lowest *P* value in the analysis of any hernia (N=11) or any hernia excluding diaphragmatic (N=28), illustrating the power gained by this approach. With MultiPhen we assessed a set of associated hernia types for each variant and broke it down to the 81 loci. The majority of 46 loci was mainly associated with inguinal hernia, but there were 26 loci associated with multiple hernias(**Figure 3**), strongly suggesting similarities in genetic mechanisms relevant for the different forms of hernia. There were clear differences by sex for the association observed at some of the loci. However, factors like age at diagnosis or different sub-diagnoses might also play a role, which were not available or only available for a subset of the data.

Genetic correlation analysis between the hernia types substantiated the connections seen in our scans at the associated variants level and also confirmed the special role of diaphragmatic hernia as three of its four correlations were at the low end of the estimates. We identified further genetic links to other traits: positive associations between diaphragmatic hernia and several anthropometric traits are also seen in other hernias, for inguinal hernia the associations are reversed, i.e. an apparently protective effect of increased BMI, which is in line with previous epidemiological studies^3,21–24^. Negative correlations with educational attainment were observed for umbilical and ventral hernia. In the psychiatric field positive associations between diaphragmatic / ventral hernia and depressive symptoms /major depression were found.

Vulnerability of connective tissue is a plausible cause for herniation of the abdominal wall. The appearance of connective tissue depends on the physical and biochemical properties of the extracellular matrix (ECM)^25^, and the ECM is composed of proteins including collagen and elastin. With this in mind we reviewed the genes in the vicinity of the identified loci with FDR below 5% in the DEPICT gene prioritization and identified several good candidates: *ADAMTS6* was already discussed in the earlier inguinal hernia GWAS^7^, due to the role of *ADAMTS* genes in encoding proteases which convert procollagen to collagen^26^, *ADAMTS16* is another member of this gene family located in the multi-variant locus on chromosome 5p15.32 (tagging variant rs7715383), and *ADAMTSL3* on 15q25.2 (tagged by the intronic variant rs551555687) is from the *ADAMTS-like* family of genes and known to play a role in ECM processes^27^. *ELN* on 7q11.23 (tagging variant rs10224499) directly encodes elastin, the protein in the ECM relevant for the elasticity of many tissues and organs. *LOX* on 5q23.2 (tagged by the intronic variant rs3835066) encodes a lysyl oxidase protein, which is important for cross-linking collagen and elastin molecules^28^. Other genes with FDR < 5% in the DEPICT gene prioritization close to identified loci were from the transforming growth factor β family (*TGFB2, BMP5, BMP7*) encoding cell regulatory proteins with many basic functions, and *LTBP1* is relevant for TGF β activity and two more genes are coding ECM proteins (*FBLN2* and *MFAP4*). Further evidence for an involvement of several of these candidates was derived by an inguinal hernia specific expression analysis resulting in nine genes meeting the significance threshold, eight of which were in loci reaching genome-wide significance (*ADAMTS6, DNAJC27, EFEMP1, FBLN2, HAND2, LOX, MFAP4*).

Overall, our study provides valuable insight into the genetics of herniation, identifying loci relevant for individual and multiple forms of hernia. Especially the fact that we identified additional loci by analyzing combined groups of hernia types can inspire future GWAS of other conditions that are also related amongst each other. The large number of associated loci for inguinal hernia point to several genes relevant for connective tissue homeostasis and general cell processes, which could be helpful to define genetic risk profiles or develop biomarkers indicating an increased vulnerability.

## Supporting information

Fadista Supplementary Material

## Data Availability

GWAS summary statistics from the UK Biobank analyses in this study will be made available via the Danish National Biobank website (https://www.danishnationalbiobank.com/gwas) upon publication of the study.

## SUPPLEMENTARY INFORMATION

### Supplementary Note contains

- Miami plots for the 7 analyzed hernia groups.
- DEPICT network plots for 36 Meta gene sets and example meta gene set embryonic morphogenesis.
- List of iPSYCH-group
- List of FinnGen contributors

**SupplementaryTable 1:** Further details on the UK Biobank and replication study groups, composition of the combined hernia groups, Pearson correlation coefficients for the 7 analyzed hernia groups, genomic control factors applied to the different SAIGE GWAS scans, hernia case codes in the UK Biobank.

**Supplementary Table 2**: Results from conditional and joined analyses of SAIGE results with GCTA-COJO at *P* < 5 × 10^−8^ in all males and females.

**SupplementaryTable 3**: Results from the inguinal hernia meta-analysis in all, males and females; collapsed loci derived from GCTA-COJO results and the inguinal hernia meta-analysis at *P* < 5 × 10^−8^; heterogeneity analysis of the collapsed loci; MultiPhen results for 81 identified loci; regression results for locus on chromosome 7q33 and potential new loci.

**SupplementaryTable 4**: Results from MAGMA gene prioritization and FUMA GTEx gene expression analyses based on complete SAIGE GWAS results.

**SupplementaryTable 5**: DEPICT results based on associated variants with *P* < 1 × 10^−5^: gene prioritization, genesets and tissue enrichment, Gene Network results for DEPICT genes.

**SupplementaryTable 6**: GARFIELD enrichment analyses (Hotspots, Peaks, Footprints, Genic, TFBS, Histone Modifications, Chromatin States and FAIRE) for the 7 analyzed hernia groups.

**Supplementary Table 7**: Bivariate LD score regression between the five different forms of hernia and 245 traits, between the hernias and six psychiatric disorders, heritability estimates for the five forms of hernia.

**SupplementaryTable 8**: PLINK clumping results for associated variants with *P* < 1 × 10^−5^ used as input for DEPICT.

## Online Methods

### Study groups

Additional details on the study groups are given in **Supplementary Table 1.**

### UK Biobank

UK Biobank is a large-scale biomedical database and research resource containing genetic, lifestyle and health information from half a million UK participants. Phenotype and imputed genotype data are available for research^8^. The questionnaire and hospital codes used for the five hernia types are listed in Supplementary Table 1. The combined groups of any hernia and any hernia excluding diaphragmatic hernia also included 4082 records of unspecific abdominal hernia repair from the interview on operations.

### FinnGen

FinnGen (https://www.finngen.fi/en) is a large biobank study that aims to genotype 500,000 Finns and combine this data with longitudinal registry data including The National Hospital Discharge Registry, Causes of Death Registry and medication reimbursement registries, all these linked by unique national personal identification codes. FinnGen includes prospective and retrospective epidemiological and disease-based cohorts as well as hospital biobank samples. The fifth data freeze (spring 2020) used in this study consists of 218,792 individuals. Inguinal hernia cases were identified via The National Hospital Discharge Registry and Causes of Death Registry.

### Estonian Biobank

Estonian Biobank samples were genotyped in Core Genotyping Lab of Institute of Genomics, University of Tartu using Illumina GSAv1.0, GSAv2.0, and GSAv2.0_EST arrays. Altogether 155,772 samples were genotyped and PLINK format files were created using Illumina GenomeStudio v2.0.4. Individuals were excluded from the analysis if their call-rate was < 95% or sex defined based on heterozygosity of X chromosome did not match sex in phenotype data. Variants were filtered by call-rate < 95% and HWE *P* value < 1e^-4^ (autosomal variants only). Variant positions were updated to build 37 and all variants were changed to be from TOP strand using tools and reference files provided in https://www.well.ox.ac.uk/~wrayner/strand/webpage. Before imputation variants with MAF<1% and indels were removed. QC was carried out using PLINK v1.9^29^ and several in-house R and PERL scripts. After QC the dataset contained 154,201 samples for imputation. Prephasing was done using Eagle v2.3^30^ and imputation was done using Beagle v.28Sep18.793^31^ with effective population size ne=20,000. Estonian population specific imputation reference of 2,297 whole genome sequencing samples were used^32^. Analyses were restricted to individuals with European ancestry. Inguinal hernia cases were identified from electronic health records (ICD-10 codes K40, K40.0, K40.1, K40.2, K40.3, K40.4 and K40.9). Association analysis of the imputed SNPs was performed with SAIGE^9^, adjusting for year of birth, sex and the first six principal components. The analysis had 3,151 cases and 138,028 controls.

### iPSYCH

The iPSYCH study group included 78,050 individuals with GWAS data from the Illumina Infinium PsychChip v1.0^20^. Data cleaning involved filtering out SNPs that had a MAF of < 0.01 or deviated from Hardy-Weinberg equilibrium (*P* < 1 × 10^−6^). Structural variants, tri-allelic SNPs, non-autosomal SNPs, and SNPs that were strand ambiguous (A/T or C/G) or did not uniquely align to the genome were also excluded, resulting in an imputation backbone of 246,369 autosomal SNPs. A total of 364 samples failing basic QC (discordant sex information, more than 1% missing genotypes, abnormal heterozygosity, duplicate sample discordance) were also excluded. Based on the good quality SNPs, all individuals were phased in a single batch using SHAPEIT3^33^ and imputed in 10 batches using Impute247 with reference haplotypes from the 1000 genomes project phase 3. Prior to analysis, we filtered out participants who were (1) of non-European ancestries, (2) related to another participant in the sample (corresponding to an IBD proportion >0.1875). We balanced the ratio of inguinal hernia cases to controls, so that it was the same within each of the iPSYCH sex-specific groups, resulting in the analysis of 2,557 cases and 35,538 controls.

### Danish diverticular disease

We used cases and controls from a Danish diverticular disease study, as described earlier^34^. Samples were genotyped with the Illumina OmniExpress-24v1-2_A1 array and we applied the following quality control steps, with PLINK v1.90b3o^29^, in order to sequentially remove (1) variant probes not uniquely aligned to the genome; (2) variants not present in dbSNP v.146, (3) variants with conflicting reference alleles between Illumina and dbSNP; (4) variants with missingness >5%, (5) variants that failed Hardy-Weinberg test at *P* < 1× 10^−6^, (6) variants with p < 0.01 in tests of differences in missingness between cases and controls; (7) variants with minor allele frequency ≤ 1%, (8) samples with discordant sex between registry records and genotypes; and (9) samples with missingness > 2%. We also removed related individuals up until third degree relatives (kinship coefficient Phi > 0.05 estimated with KING^35^ as implemented in VCFtools^36^). European ancestry outliers were removed based on principal component analysis of all our cases and controls combined with the European samples from the Human Genome Diversity Project^37^ using the R package SNPRelate v1.2.0^38^. Outliers were defined based on the first two principal components, excluding samples where the distance between at least one principal component and its mean was larger than 6 times the interquartile range. This outlier removal procedure was repeated until no more samples fulfilled the criterion for removal. After these steps, we imputed unobserved genotypes with phased haplotypes from the Haplotype Reference Consortium panel version r1.1^39^ on the imputation server at the Wellcome Trust Sanger Institute (https://imputation.sanger.ac.uk/). Inguinal hernia cases were defined as having an ICD-10 code K40 (and sub-codes) and/or ICD-8 codes 550 and/or 552 (and-sub-codes). Association analysis of imputed SNPs was performed with SAIGE^9^, adjusting for year of birth, sex and diverticular disease status. We had 999 inguinal hernia cases and 7768 controls probed at 6,597,770 SNPs.

### Ethics statement

Patients and control subjects in FinnGen provided informed consent for biobank research, based on the Finnish Biobank Act. Alternatively, separate research cohorts, collected prior the Finnish Biobank Act came into effect (in September 2013) and start of FinnGen (August 2017), were collected based on study-specific consents and later transferred to the Finnish biobanks after approval by Fimea, the National Supervisory Authority for Welfare and Health. Recruitment protocols followed the biobank protocols approved by Fimea. The Coordinating Ethics Committee of the Hospital District of Helsinki and Uusimaa (HUS) approved the FinnGen study protocol Nr HUS/990/2017. The FinnGen study is approved by Finnish Institute for Health and Welfare (permit numbers: THL/2031/6.02.00/2017, THL/1101/5.05.00/2017, THL/341/6.02.00/2018, THL/2222/6.02.00/2018, THL/283/6.02.00/2019, THL/1721/5.05.00/2019, THL/1524/5.05.00/2020, and THL/2364/14.02/2020), Digital and population data service agency (permit numbers: VRK43431/2017-3, VRK/6909/2018-3, VRK/4415/2019-3), the Social Insurance Institution (permit numbers: KELA 58/522/2017, KELA 131/522/2018, KELA 70/522/2019, KELA 98/522/2019, KELA 138/522/2019, KELA 2/522/2020, KELA 16/522/2020 and Statistics Finland (permit numbers: TK-53-1041-17 and TK-53-90-20). The Biobank Access Decisions for FinnGen samples and data utilized in FinnGen Data Freeze 6 include: THL Biobank BB2017_55, BB2017_111, BB2018_19, BB_2018_34, BB_2018_67, BB2018_71, BB2019_7, BB2019_8, BB2019_26, BB2020_1, Finnish Red Cross Blood Service Biobank 7.12.2017, Helsinki Biobank HUS/359/2017, Auria Biobank AB17-5154, Biobank Borealis of Northern Finland_2017_1013, Biobank of Eastern Finland 1186/2018, Finnish Clinical Biobank Tampere MH0004, Central Finland Biobank 1-2017, and Terveystalo Biobank STB 2018001.

The use of the Estonian Biobank data in this study was approved by the Research Ethics Committee of the University of Tartu (Approval number 288/M-18).

The Danish Scientific Ethics Committee, the Danish Data Protection Agency and the Danish Neonatal Screening Biobank Steering Committee approved the iPSYCH study.

The Danish diverticular disease study was approved by the Scientific Ethics Committee of the Capital Region of Denmark (H-16016406) and the Danish Data Protection Agency. The Scientific Ethics Committee granted exemption from obtaining informed consent from participants as the study was based on the biobank material.

### Association analyses

Initial genome-wide association studies for the 7 hernia groups were performed with SAIGE^9^, a method allowing for related samples and thus maximizing power. Covariates were sex, year of birth and the first six principal components calculated for the analyzed subset of the UK Biobank with British ancestry, *P* values were adjusted by genomic control (**Supplementary Table 1**). To identify the most likely causal variants in the different GWAS scans, conditional and joined analyses^10^ of the initial SAIGE results were performed with GCTA^40^ using the COJO option. We performed these analyses with thresholds of 5 × 10^−8^ and 1 × 10^−5^ to identify variants reaching genome-wide significance for the specific hernia group and to define a larger list of potentially associated variants for subsequent analyses, respectively. The meta-analysis for the inguinal hernia replication study and the heterogeneity analyses by sex for the five individual hernia types were performed with METAL^11^.

We applied MultiPhen^12^ (a method that models multiple phenotypes simultaneously by taking the genotype as dependent variable and performing ordinal regression) to a subset of 335,258 unrelated individuals (no third degree or closer relationship). The outcome is a linear combination of the five hernia types most strongly associated with the genotypes of a variant and we selected a *P* threshold of 1% to define association for one of the five hernia groups. Potential new loci identified by MultiPhen were confirmed by linear regression models in R^13^ with the allele count at the respective variant as outcome and the identified forms of hernia as explanatory variables (both *P* < 5 × 10^−8^ for the joint models).

### Pathway and enrichment analyses

We applied several software packages to the association results to identify associated genes, genesets and pathways.

We performed MAGMA gene prioritization^14^ as implemented in FUMA^15^ based on the complete GWAS scan results, and FUMA expression results for 54 GTEx tissues were derived from MAGMA analysis.

We applied DEPICT^16^ (Data-driven Expression Prioritized Integration for Complex Traits), which has three features: gene prioritization, pathway analysis and tissue/cell type enrichment analysis. Variants from the HLA region and chromosome X were not considered by the DEPICT analyses. To provide independent loci as input, we clumped variants with *P* < 1 × 10^−5^ in PLINK (**SupplementaryTable 8**).

GeneNetwork^17^ uses 31,499 public RNA-seq sample to prioritize genes based on human phenotype ontology (HPO) terms. We generated GeneNetwork results for the HPO term inguinal hernia for the 192 genes studied in the DEPICT inguinal hernia gene prioritization.

GARFIELD^18^ (GWAS analysis of regulatory or functional information enrichment with LD correction) investigates regulatory and functional implications of GWAS findings. To focus the analyses on the variants identified in the conditional and joint GCTA-COJO analysis at P < 1 × 10^−5^, we removed all other variants with P < 1 × 10^−5^ from the input. The annotation step in GARFIELD still included all correlated variants at *r*^*2*^ > 0.8 for the variants identified by GCTA-COJO.

### Heritability and genetic correlation analysis

Heritability was estimated for the five hernia types by applying LD score regression^41,42^ to the UK Biobank discovery GWAS scans. Heritability was transformed to the liability scale with GEAR (https://sourceforge.net/p/gbchen/wiki/Hong%20Lee’s%20Transformation%20for%20Heritability/). We investigated genetic correlation between the five hernia types as well as between them and 245 traits with a PubMed ID available in LD hub^43^ by performing LD score regression.

### URLs

UK Biobank website: www.ukbiobank.ac.uk

FinnGen website: https://www.finngen.fi/en hernia phenotypes in mice from Mammalian Phenotype Ontology Annotations: www.informatics.jax.org/vocab/mp_ontology

Rayner strand alignment tool: www.well.ox.ac.uk/~wrayner/strand

Imputation server at the Wellcome Trust Sanger Institute: https://imputation.sanger.ac.uk

GEAR heritability: https://sourceforge.net/p/gbchen/wiki/Hong%20Lee’s%20Transformation%20for%20Heritability/

## Acknowledgments

We thank all participants and staff of the UK, FinnGen and Estonian biobanks and the iPSYCH and Danish diverticular disease study for their contribution to this research.

This research has been conducted using data from UK Biobank, a major biomedical database (project ID 33395). The study was supported by grants from the Oak Foundation (OCAY-18-598), a Lundbeck Foundation Ascending Investigator grant (R313-2019-554) to B.F. The Danish National Biobank was established with the support of major grants from the Novo Nordisk Foundation, the Danish Medical Research Council and the Lundbeck Foundation. L.S. received support from a Carlsberg Foundation postdoctoral fellowship (CF15-0899).

The FinnGen project is funded by two grants from Business Finland (HUS 4685/31/2016 and UH 4386/31/2016) and the following industry partners: AbbVie Inc., AstraZeneca UK Ltd, Biogen MA Inc., Celgene Corporation, Celgene International II Sàrl, Genentech Inc., Merck Sharp & Dohme Corp, Pfizer Inc., GlaxoSmithKline Intellectual Property Development Ltd., Sanofi US Services Inc., Maze Therapeutics Inc., Janssen Biotech Inc, and Novartis AG. Following biobanks are acknowledged for delivering biobank samples to FinnGen: Auria Biobank (www.auria.fi/biopankki), THL Biobank (www.thl.fi/biobank), Helsinki Biobank (www.helsinginbiopankki.fi), Biobank Borealis of Northern Finland (https://www.ppshp.fi/Tutkimus-ja-opetus/Biopankki/Pages/Biobank-Borealis-briefly-in-English.aspx), Finnish Clinical Biobank Tampere (www.tays.fi/en-US/Research_and_development/Finnish_Clinical_Biobank_Tampere), Biobank of Eastern Finland (www.ita-suomenbiopankki.fi/en), Central Finland Biobank (www.ksshp.fi/fi-FI/Potilaalle/Biopankki), Finnish Red Cross Blood Service Biobank (www.veripalvelu.fi/verenluovutus/biopankkitoiminta) and Terveystalo Biobank (www.terveystalo.com/fi/Yritystietoa/Terveystalo-Biopankki/Biopankki/). All Finnish Biobanks are members of BBMRI.fi infrastructure (www.bbmri.fi) and FinBB (https://finbb.fi/).

The work of Estonian Genome Center, Univ. of Tartu has been supported by the European Regional Development Fund and grants No. GENTRANSMED (2014-2020.4.01.15-0012), MOBERA5 (Norface Network project no 462.16.107) and 2014-2020.4.01.16-0125. This study was also funded by the European Union through Horizon 2020 research and innovation programme under grant no 810645 and through the European Regional Development Fund project no. MOBEC008 and Estonian Research Council Grant PUT1660. Data analyses with Estonian datasets were carried out in part in the High-Performance Computing Center of University of Tartu.

The content is solely the responsibility of the authors and does not necessarily represent the official views of the funders.

## Author contributions

F.G. conceptualized the study and wrote the original manuscript draft. J.F. and F.G. coordinated the analyses. J.F., L.S., J.K., E.A. and F.G. analyzed the data. E.S. and H.U. provided data from the diverticular study. T.W. provided data from the iPSYCH study. T.E. and L.M. provided data from the Estonian Biobank study. A.P. and M.D. provided data from the FinnGen study. J.F., L.S., M.M., B.F. and F.G. interpreted the results. M.M., B.F. and F.G. supervised the study. All authors contributed to the revision of the manuscript and approved it for submission.

## Competing interests

J.F. currently works for Novo Nordisk A/S.

## Notes

### Competing Interest Statement

Joao Fadista currently works for Novo Nordisk A/S.

### Author Declarations

Patients and control subjects in FinnGen provided informed consent for biobank research, based on the Finnish Biobank Act. Alternatively, separate research cohorts, collected prior the Finnish Biobank Act came into effect (in September 2013) and start of FinnGen (August 2017), were collected based on study-specific consents and later transferred to the Finnish biobanks after approval by Fimea, the National Supervisory Authority for Welfare and Health. Recruitment protocols followed the biobank protocols approved by Fimea. The Coordinating Ethics Committee of the Hospital District of Helsinki and Uusimaa (HUS) approved the FinnGen study protocol Nr HUS/990/2017. The FinnGen study is approved by Finnish Institute for Health and Welfare (permit numbers: THL/2031/6.02.00/2017, THL/1101/5.05.00/2017, THL/341/6.02.00/2018, THL/2222/6.02.00/2018, THL/283/6.02.00/2019, THL/1721/5.05.00/2019, THL/1524/5.05.00/2020, and THL/2364/14.02/2020), Digital and population data service agency (permit numbers: VRK43431/2017-3, VRK/6909/2018-3, VRK/4415/2019-3), the Social Insurance Institution (permit numbers: KELA 58/522/2017, KELA 131/522/2018, KELA 70/522/2019, KELA 98/522/2019, KELA 138/522/2019, KELA 2/522/2020, KELA 16/522/2020 and Statistics Finland (permit numbers: TK-53-1041-17 and TK-53-90-20). The Biobank Access Decisions for FinnGen samples and data utilized in FinnGen Data Freeze 6 include: THL Biobank BB2017_55, BB2017_111, BB2018_19, BB_2018_34, BB_2018_67, BB2018_71, BB2019_7, BB2019_8, BB2019_26, BB2020_1, Finnish Red Cross Blood Service Biobank 7.12.2017, Helsinki Biobank HUS/359/2017, Auria Biobank AB17-5154, Biobank Borealis of Northern Finland_2017_1013, Biobank of Eastern Finland 1186/2018, Finnish Clinical Biobank Tampere MH0004, Central Finland Biobank 1-2017, and Terveystalo Biobank STB 2018001. The use of the Estonian Biobank data in this study was approved by the Research Ethics Committee of the University of Tartu (Approval number 288/M-18). The Danish Scientific Ethics Committee, the Danish Data Protection Agency and the Danish Neonatal Screening Biobank Steering Committee approved the iPSYCH study. The Danish diverticular disease study was approved by the Scientific Ethics Committee of the Capital Region of Denmark (H-16016406) and the Danish Data Protection Agency. The Scientific Ethics Committee granted exemption from obtaining informed consent from participants as the study was based on the biobank material.

